# Spread of the Novel Coronavirus (SARS-CoV-2): Modeling and Simulation of Control Strategies

**DOI:** 10.1101/2020.05.11.20098418

**Authors:** Harishankar Prabhakaran

## Abstract

The coronavirus disease 2019 (COVID-19) is spreading throughout the world and all healthcare systems are loaded beyond its capacity. The virus is named as SARS-CoV-2. In this situation, rational decisions need to be made on how the care is provided for patients with COVID-19. The Incidence report, general symptoms and readily available testing kits, different control strategies, the basic compartmental model, and some of the current research on the epidemiology of the disease are discussed and previously published models are reviewed. Modeling this disease helps in understanding the spread, and predict its future to evaluate different control strategies (Social Distancing, Contact Tracing and Hospitalization). Compartmental modeling framework is used in this work. The non-linear equations are formulated and fitted to the cumulative case and mortality data. Analytical analysis along with uncertainty analysis and sensitivity analysis is performed, and the conditions to achieve disease free equilibrium is evaluated. Finally, Different control strategies are simulated to show their importance. This paper aims to shows the advantage of mathematical modeling and their simulations in times like now, during which the COVID-19 spreading like wildfire. It also includes Pre-symptomatic and asymptomatic individuals in the modeling. The simulations are performed for the model fit to Cumulative Case and Mortality data in the United States of America. The Reproduction number is found to be **2.71914**.

## 1 Introduction

This paper is an effort to model the epidemiology and stop the spread of the novel coronavirus, which is causing huge detrimental effects all around the world. The ultimate goal is to slow the spread of the virus, using evidence based public health interventions, which includes early case recognition, isolation and contact tracing. This disease has grown from being a small cluster of pneumonia in china to a global pandemic causing huge loss of life. Therefore, there is a huge need for modeling the disease, and simulation of various control strategy implementation is necessary to make well informed decisions.

### 1.1 Incidence

Incidence is the number of new cases of coronavirus in a particular population in a period of time. In general, incidence is reported for each country’s population. The table below gives the incidence of all the country that has been affected majorly.

**Table 1:** Characteristics of countries with Majority of cases, Disease Incidence and Mortality of coronavirus disease 2019 (COVID-19) as of 27 April 2020 [1]

| Country<br>(Population in Millions)<br>till April 27<br>(Data) | Cumulative<br>Number<br>of Cases<br>(Data) | Number<br>of<br>Deaths<br>(Data) | Mortality<br>Rate (%)<br>(CFR)<br>(Calculated) | Incidence<br>per<br>1Million<br>(Calculated) | Deaths<br>per<br>1Million<br>(Calculated) |
| --- | --- | --- | --- | --- | --- |
| Western Pacific Region |  |  |  |  |  |
| China (1393) | 84341 | 4643 | 05.5050 | 60.54 | 03.33 |
| Singapore (5.639) | 13624 | 12 | 00.0880 | 2416.03 | 02.12 |
| Japan (126.5) | 13385 | 351 | 02.6223 | 105.81 | 02.77 |
| European Region |  |  |  |  |  |
| Spain (46.94) | 207634 | 23190 | 11.1686 | 4423.39 | 494.03 |
| Italy (60.36) | 197675 | 26644 | 13.4786 | 3274.93 | 441.41 |
| Germany (83.02) | 155193 | 5750 | 03.7050 | 1869.34 | 069.26 |
| The United Kingdom (66.65) | 152844 | 20732 | 13.5641 | 2293.23 | 311.05 |
| France (66.99) | 123279 | 22821 | 18.5116 | 1840.25 | 340.66 |
| South-East Asia Region |  |  |  |  |  |
| India (1353) | 27892 | 872 | 03.1263 | 20.61 | 0.64 |
| Eastern Mediterranean Region |  |  |  |  |  |
| Iran (81.8) | 90481 | 5710 | 06.3107 | 1106.12 | 69.80 |
| Saudi Arabia (33.7) | 17522 | 139 | 00.7932 | 519.94 | 04.12 |
| Pakistan (212.2) | 13328 | 281 | 02.1083 | 62.80 | 01.32 |
| Region of the Americas |  |  |  |  |  |
| USA (328.2) | 931698 | 47980 | 05.1497 | 2838.81 | 146.19 |
| Brazil (209.5) | 58509 | 4016 | 06.8639 | 279.27 | 19.16 |
| Canada (37.59) | 45778 | 2489 | 05.4371 | 1217.82 | 66.21 |
| African Region |  |  |  |  |  |
| South Africa (57.78) | 4546 | 87 | 01.9137 | 78.67 | 01.50 |
| Algeria (42.23) | 3382 | 425 | 12.5665 | 80.08 | 10.06 |
| Global - All Regions |  |  |  |  |  |
| Whole World (7594) | 2878196 | 198668 | 06.9025 | 379.00 | 26.16 |

The incidence per 1Million calculated in the Table gives a better picture than the number of cases, on how the country is affected. For example, by comparing India and Singapore, India has twice the number of cases than in Singapore, and it does not mean that India more is affected. The incidence per 1 Million for Singapore is more than 100 time to that of India’s. It is because the population of Singapore is far less than India.

The death per 1Million data also gives the information on how each country is handling the infected cases.

Among all the countries, Spain is in a very bad state with a mortality rate of 11.16%. The United States of America has the most number of cases (1.2 Million as of May 8th, 2020) and has more than 75,000 deaths.

### 1.2 Spread of Virus from one to another

The Coronavirus spreads mainly from person to person, through respiratory droplets from an infected person (via. coughs or sneezes). These droplets can go into people who are nearby. It spreads, when people are near to each other. The droplets do not hang around in the air and can not go more than six feet from the source (about two meters).

COVID-19 spreads sustainably in a community in many affected Regions. People are getting infected with the virus, mostly without knowing where they are infected.

People also get infected when they come in contact with an infected surface and then touches their eyes or face.

### 1.3 Symptoms and Testing

Symptoms for COVID-19 is not constant, and ranges from mild disease to severe illness. Symptoms usually appears 14 days after being exposed to the virus. Major indications of COVID-19 are Cough, Shortness of breath or difficulty breathing, Fever, Chills, Muscle pain, Sore throat, New loss of taste or smell [2].

Medical attention should be sought immediately if the following symptoms are noticed: Trouble breathing, Persistent pain or pressure in the chest, New confusion or inability to arouse, Bluish lips or face [2].

With the restricted availability of testing kits, not everyone is eligible for being tested. If it only causes mild illness, people are asked to recover at their home itself. Decisions on who to be tested are made by health departments and healthcare providers.

By the end of April, a viral test has been authorized by the U.S. Food and Drug Administration (FDA) that lets people to collect a respiratory sample (a swab inside of the nose) at home, and send it to a laboratory for analysis. In most areas the collection kits were made available via healthcare workers [3]..

### 1.4 Control Strategies

#### 1.4.1 Social Distancing

Things to do to implement social distancing: Avoid contact with people who could be at risk, Avoid public places, gathering and crowds, Avoid clinics and hospitals, and Avoid restaurants.

It is every individuals responsibility to prevent this virus from spreading, and social distancing is a way collective effort in stopping the spread of this disease. In case of emergencies in which we can not avoid going to different places, one should maintain at least a distance of 6 feet from one another.

#### 1.4.2 Contact Tracing

Contact tracing is carried out by a public health staff. They get data from the patients with coronavirus to help everyone they have had contacted during the previous weeks while they were infectious. The identity of the patient is not revealed for their privacy. Traced individuals are given information on the risk of being infected, and are asked to separate themselves from other susceptible people, and check for symptoms and illness. They are asked to monitor their body temperature twice a day, and if they have symptoms, they should isolate themselves and report to a health care worker.

#### 1.4.3 Hospitalization/Isolation

Hospitalization or isolation is the process of separating the sick people from the non sick ones. With a steady rise in new cases of coronavirus, hospitals are working at maximum capacity, and infected individuals are admitted only when they show severe symptoms or have extreme illness. Temporary medical clinics can be setup in areas where most infections occur can reduce the spread of the disease.

#### 1.4.4 Quarantine

Quarantines are used by governments to prevent the spread of the virus. It is especially for individuals have no symptoms but were exposed to the sickness. It is important to keep them separated from susceptible people so that they don’t infect others without their knowledge. Quarantines are used during an outbreak, or an epidemic, or during a pandemic.

#### 1.4.5 Masks

It has been debated whether masks are effective or not. The truth is that it works. The effect of face masks is illustrated very clearly in the paper, ‘To mask or not to mask’ [14].

Both hands should be cleaned before wearing the mask. There should be no gaps between ones face and the mask. Never touch the mask, and discard them in a closed bin quickly, once they are damp. Masks are effective only if the hands are washed properly and the used masks are disposed properly.

#### 1.4.6 Hand washing

Hands should be cleaned with alcohol-based hand rub or soap and water. Our hands mostly touch different surfaces when we go out. Virus sticks to ones hands and can infect us. Therefore, it is important to wash our hands regularly to prevent the infection.

### 1.5 Risk-High groups

Elderly people are affected the most compared to the young population. Mortality rate for people in 75 to 84 years is in the range of 4-10%, and the rate goes up to an average 20% for people who are more than 85 years old. [22]

Individuals with medical conditions like heart/lung disease or diabetes are at a greater risk for developing serious illness due to the disease [4]. People who are most affected are ones with the following conditions: asthma, immunocompromised conditions, severe obesity, chronic kidney disease undergoing dialysis, and liver diseases.

#### 1.6 Kermack-Mckendrick model

The Kermack-McKendrick model [5] is a SIR epidemiological model. It explains the rapid rise and fall in the number of infected cases reported in an epidemic. The model assumes that there is no change in the total population size (i.e., no vital dynamics and deaths due to disease), and that there is no incubation period.

The model has three nonlinear ODEs:

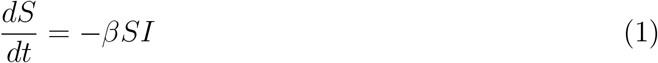

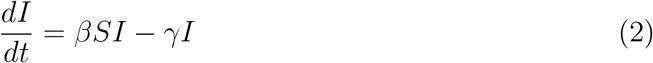

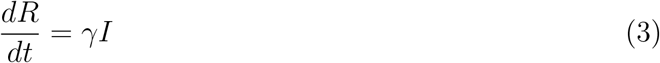

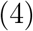

where, S is the number of susceptible people, I is the number of Infected people, R is the number of recovered people who gained immunity, *β* is the infection rate, and *γ* is the recovery rate.

The Reproduction number of this model is was called as epidemiological threshold.

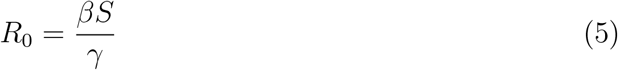

To control the disease outbreak, *R*_0_ should be less than 1.

This basic model structure first published in 1927 have been developed into many complex epidemiological models over a century, and currently there are thousands of published models. This paper also uses the same type of modeling methodology.

### 1.7 Literature Review of Major Models of COVID-19

As there are many uncertainties in hoe the corona-virus is spreading, there is is a huge need for modeling the spread of the disease. Many papers are being published once the pandemic began. The most predominant modeling papers are discussed here.

- A mathematical model for simulating the transmission of Wuhan novel Coronavirus [6] is a different Paper than other modeling papers being published. They have developed a Bats-Hosts-Reservoir-People transmission network model. They have simplified their model as Reservoir-People transmission network. It is a brief paper with the modeling framework, the nonlinear equations and the first generation method derivation for the reproduction number.
- A mathematical model for the novel coronavirus epidemic in Wuhan, China [7]. The model is a basic SEIR compartments with a V compartment(concentration of the coronavirus in the environmental reservoir) at the end. The model also includes the vital dynamics of the population. They got a high Reproduction number estimate of 4.25. The inclusion of the vital dynamics in the model seems to have increased the ability of the disease to spread more. It is also noticeable that they have modeled three different transmission rate for every class that can be infectious. The paper is concentrated towards Wuhan and therefore the dynamics vary a little from other modeling papers.
- Report 9: Impact of non-pharmaceutical interventions (NPIs) to reduce COVID19 mortality and healthcare demand.[16] This report is a very important, because many policies in the UK and other countries are made based on it. It discusses about various non-pharmaceutical interventions that are aimed at reducing contact rates in the population, and the conclusion was that no intervention can impact the contact rate if they are implemented separately. All interventions should be implemented to achieve a synergistic effect.
- Forecasting the impact of the first wave of the COVID-19 1 pandemic on hospital demand and deaths for the USA and 2 European Economic Area countries.[8]. This paper used data identification tools and statistical modeling to model the disease. They have predicted a huge demand for hospital and ICU bed-days, and their estimation of the number of deaths of around 1,584,737 in USA from the first wave of seems unlikely. This gives us an idea on how compartmental modeling is better than statistical data analysis for epidemiological models.
- Flattening the curve before it flattens us: hospital critical care capacity limits and mortality from novel coronavirus (SARS-CoV2) cases in US counties. [9]. This paper concentrates on the relation between available critical care and deaths that occurs when critical care bed limits are exceeded. They have devised for 4 critical care surge response scenarios. Their concern is that the mortality data does not include normal patients with injury, who could not get treated due to suddenly overloaded healthcare system.
- To mask or not to mask: Modeling the potential for face maskuse by the general public to curtail the COVID-19 pandemic. [14]. This paper considers the importance of face mask in the prevention of the disease. Two models, one with no mask use and the other with mask use and control strategies are created. Simulations are done in both these models and the results are compared. It talks about inward efficiency (prevention from catching the disease) and outward efficiency (preventing the transmission of the disease). The model for the exponentially decaying infectious contact rate in this work is inspired from this paper. The model is parameterized for the data for New York and Washington. They found that the usage of masks have a huge impact than what we already believed it to have.
- A Case Study of the COVID-19 Coronavirus. Mathematical Modeling of Epidemic Diseases. [10]. This paper deals with simple models, but demonstrated the effects of various control strategies. It begins with an explanation of the basic SIR model, followed by compartmental SEIR models for the coronavirus. The paper also compares two scenarios where in one, the recovered people gets lifetime immunity, and the other has re-Susceptible factor in the model. Other simulations such as short lock down, and healthcare system saturation is also presented.
- REFINED COMPARTMENTAL MODELS, ASYMPTOMATIC CARRIERS AND COVID-19. [11]. This paper talks about the various possible compartmental models which can be used as the base model for the novel coronavirus. A discussion on these models (modified SEIR, splitting the exposed group into different compartments, and including asymptomatic population), explains them well. This paper is highly illustrative with many simulations.
- Construction of Compartmental Models for COVID-19 with Quarantine, Lock-down and Vaccine Interventions [12]. Many epidemiological models are formed and the dynamics are analysed. They also explore the transmissibility of the viral infection during different control strategies like quarantine, lock-downs, and vaccinations. The dynamics of coronavirus is simulated for the data in Ghana. It is interesting to see that the basic reproduction number is less than 1, and they expect that the disease will be gone in two to four months. The graphical representation of the model framework used in this work is inspired from this paper.
- Modeling the Spread of COVID-19 with COMSOL Multiphysics [13]. COMSOL Multiphysics is a virtual dynamical simulation software. They have used the software to simulate the compartmental models with an aim to flatten the curve. They have presented a basic SEIR model an a SEIR model with many sub-compartments of Exposed and Infectious class. It is amazing how a physics simulation software can be used to simulate dynamics of epidemiology.

## 2 Methods

### 2.1 Modelling the framework & finding parameter values

#### 2.1.1 S-E-P-I-A-H-C-R-D Model Definition

A deterministic model of the Coronavirus 2019 disease transmission, is used to estimate the parameters in The United States of America. This model includes factors which reduces the transmission of the disease, such as social distancing, Hospitalization and Contact tracing of Asymptotic Individuals.

The modeling framework consists of susceptible, exposed, pre-symptomatic infectious, symptomatic infectious, asymptomatic infectious, hospitalized-non ICU, hospitalized ICU,and recovered, with the classes respectively denoted S(t), E(t), P(t), I(t), A(t), H(t), C(t), and R(t). In addition, a cumulative death class is also included and is denoted as D(t).

After the incubation period in exposed class, they become infectious, but they are pre-symptomatic for a period; a part of them eventually become symptomatic and the others asymptomatic. A fraction of detected infectious individuals (by symptoms for I(t) class; by contact tracing for A(t) class) are moved to the hospitalized class (non-ICU), H(t), where they are isolated from general public. Some fraction of hospitalized patients ultimately require critical care and they are moved to hospitalized ICU class. Some fraction recover and some die in the classes I(t), A(t), H(t) and C(t). Based on these assumptions, the deterministic model for the transmission dynamics of COVID-19 is given by the framework in Figure 1, and the corresponding deterministic system of nonlinear differential equations is given below.

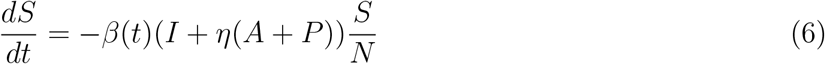

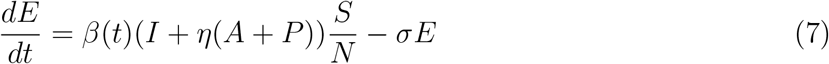

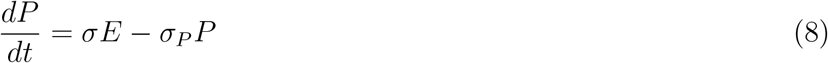

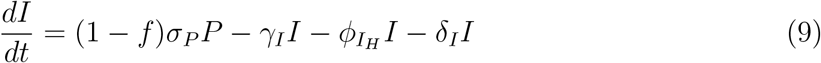

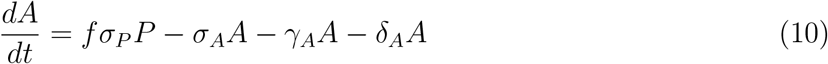

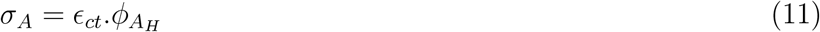

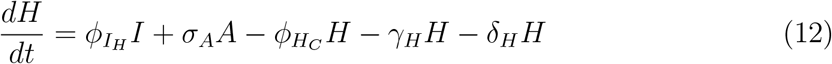

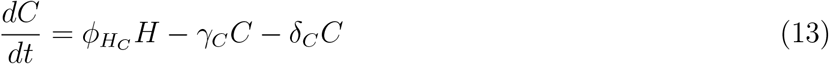

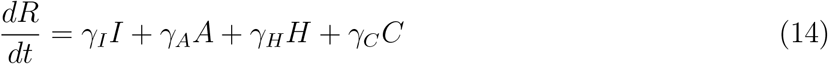

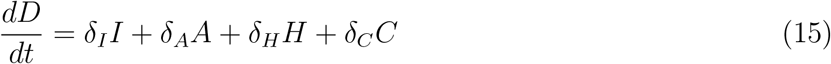

where, N is the total population.

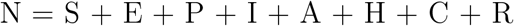

**Figure 1:**
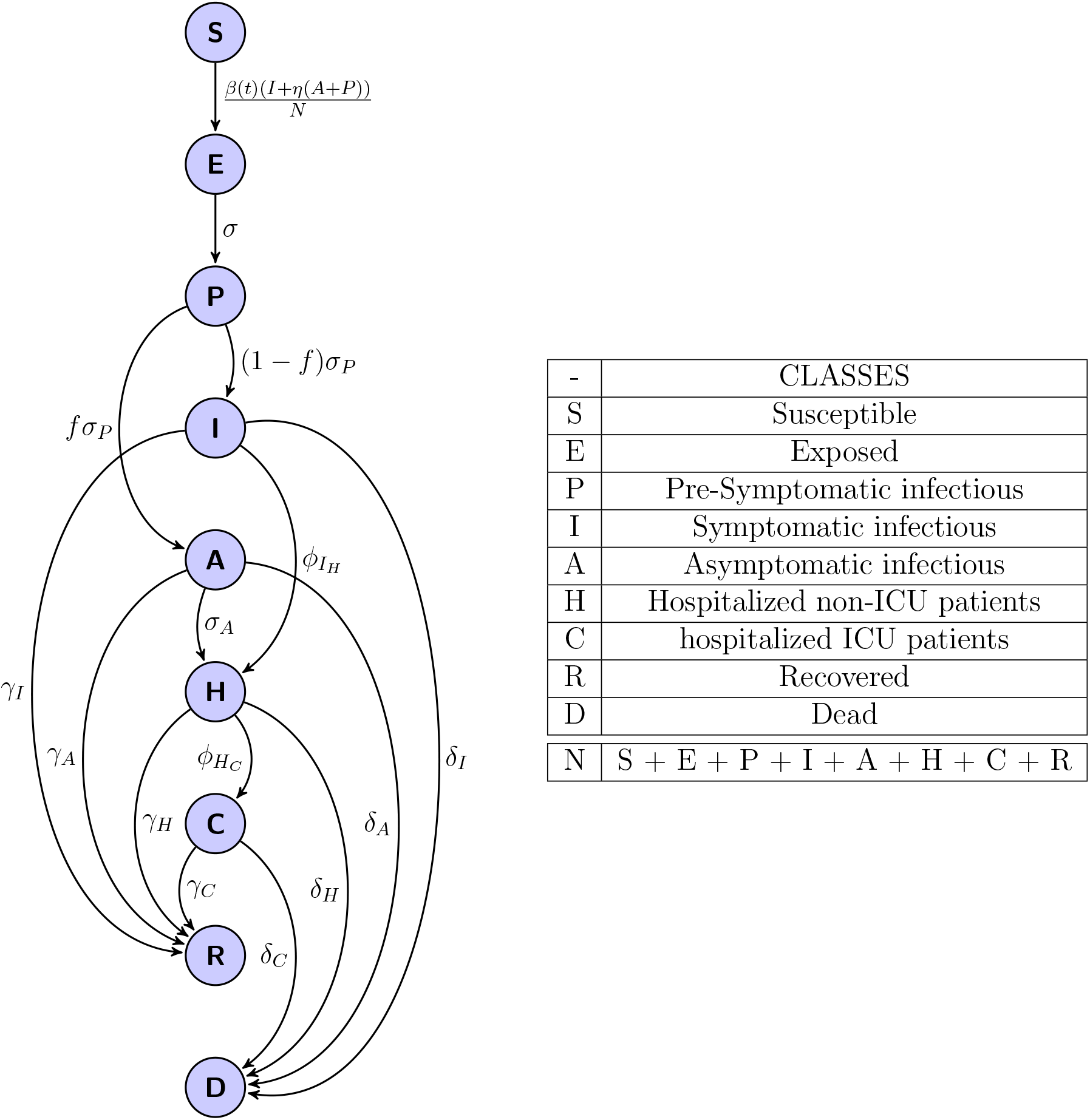
The model Framework

#### 2.1.2 Model Parameters

All the parameters in the model is defined and a default value is estimated. They are found by searching research literature, official reports or by fitting the model to the available data.

*β*(t) is the time dependent infectious contact rate, which has a exponentially decreasing structure [14]. The decrease is due to the prevention measures put into action by the government and good will of the people.

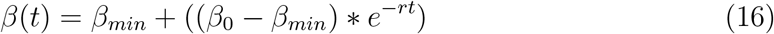

We assume that *β* decrease right from the beginning (without delay) because our initial condition *β* _0_ for the model is set to be on 01-March-2020, during when they started to implement preventive measures. It decreases to *β_min_* as the time passes at the rate of *r*. *β_min_* required to get to disease free equilibrium is calculated from conditions for the basic reproduction number. The *β_min_* and *r* are fixed during the model fitting process.

**Table 2:**
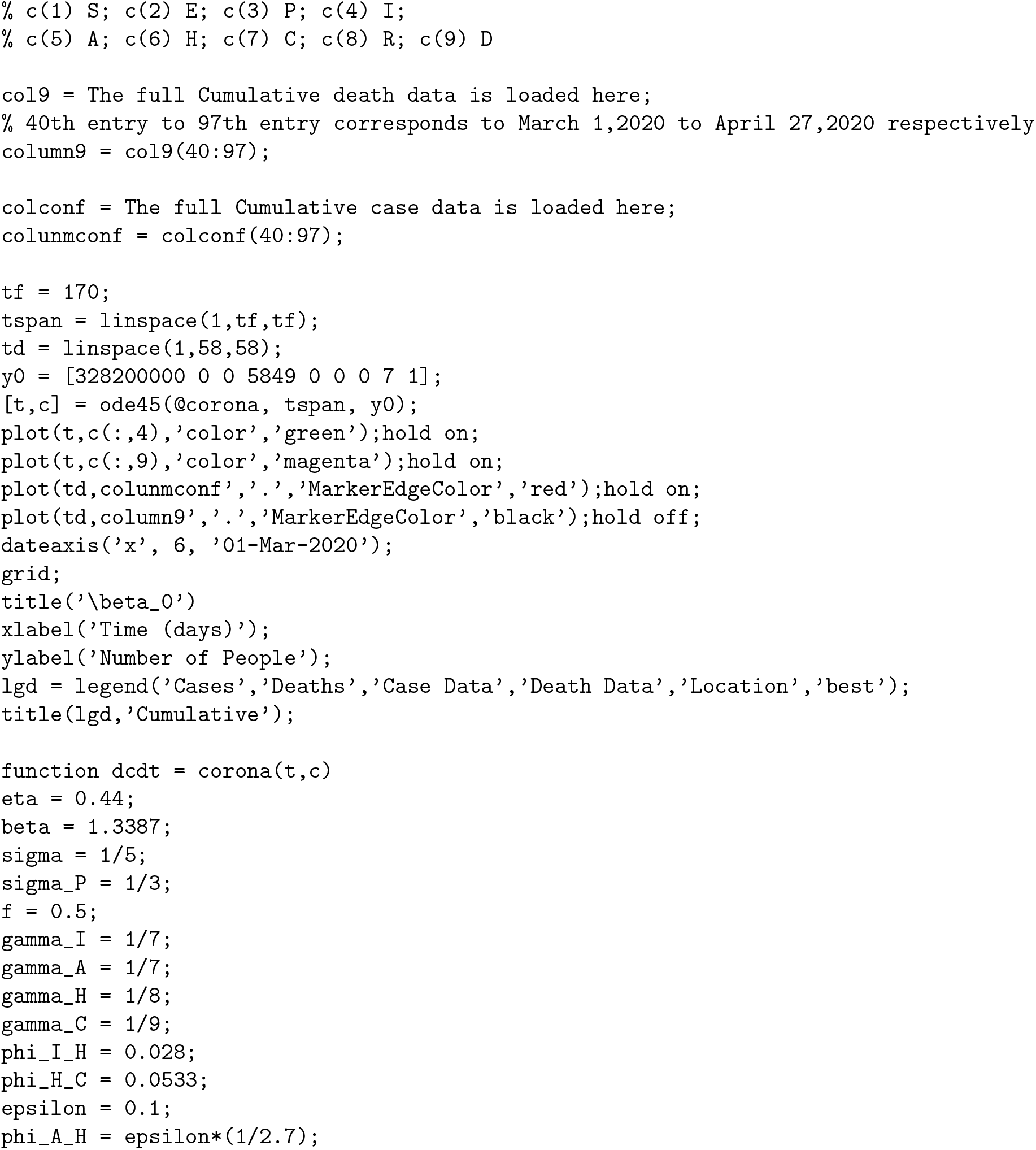
Model Parameters

- *η* the relative infectiousness of asymptomatic carriers compared to symptomatic carriers, and it is assumed that symptomatic individuals are 50% more infectious than the asymptomatic people [16]. The value is set to 0.44.
- *σ* is the transition from exposed to pre-symptomatic class, where 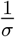 is the incubation period (average 5 days [17] for Coronavirus). So, the value of *σ* is 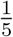.
- *σ_P_* is the transition from pre-symptomatic to infectious class (I or A)). Calculated similar to *σ*. The period of stay in this class is an average 3 days. *”This virus continues to spread across the country, because we have asymptomatic transmitters and we have individuals who are transmitting 48 hours before they become symptomatic”* - CDC Director [19].
- *f* is the fraction of pre-symptomatic individuals who eventually becomes asymptomatic. There is a lot of uncertainty in this fraction [20]. The average value is set to be 0.5.
- 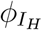 is the rate of hospitalization of symptomatic people. The median interval from symptom onset to admission was 7 days (interquartile range [IQR] = 3−9 days)[21]. The percentage of hospitalization is 20 [22]. So, 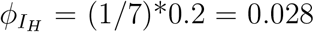.
- *σ_A_* is the rate at which asymptotic individuals are detected using contact tracing and hospitalized. It is the product of the percentage of contact-tracing execution and the rate of asymptomatic hospitalization. Contact tracing reduced the time taken to identify and isolate patients after onset of symptoms to 2.7 days [23]. *ϵ_ct_* ia control variable (varied in the simulations) 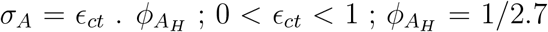 Default *ϵ_ct_* is taken to be 0.1. Therefore, default *σ_A_* is (1/2.7)*0.1 = 0.037.
- 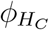 is the rate of ICU admission from Hospitalization. In a study by Chen Wang et al. [25], of the patients hospitalised 32% required admission to an intensive care unit. In another report from Imperial College [24], they calculate bed demand numbers assuming a total duration of stay in hospital of 8 days if critical care is not required and 16 days (with 10 days in ICU) if critical care is required. With 30% of hospitalised cases requiring criticalcare, they obtained an overall mean duration of hospitalisation of 10.4days, slightly shorter than the duration from hospital admission to discharge observed for COVID-19 cases internationally. Therefore, if someone needs critical care, on an average they stay in the hospital for 6 days before moving to ICU. So, 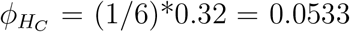. Also *γ_H_*, the recovery rate of Hospitalized people for whom critical care is not required is 1/8; and *γ_C_*, the recovery rate of Hospitalized in need of ICU is 1/9 [26].
- Both *γ_i_*, the recovery rate of Symptomatic individuals and *γ*_A_, the recovery rate of Asymptomatic individuals, is on an average found to be 1/7. It is because, on an average the people in these classes recover in 7 days.
- *δ_I_*, the death rate of Symptomatic individuals. It is found that the average duration between the onset of symptoms and death is 22 days [26]. The mortality rate among the cases found is 3.4% [28]. So, *δ_I_* = (1/22)*0.034 = 0.00154.
- *δ_a_* is the death rate of the Asymptomatic population, is assumed to be half of *δ_I_* [29] So, *δa* = *δ_I_* / 2 = 0.00077.
- *δ_h_* (death rate, Hospitalized - Non ICU) It is found that the average duration between hospitalization and death is 8 days [26]. With the mortality rate of 3.4%, *δ_h_* = (1/8)*0.034 = 0.00425.

### 2.2 Model fitting

The state-level time series for both cumulative mortality data and cumulative case data, compiled by Center for Systems Science and Engineering at Johns Hopkins University [31], from March 1, 2020, through April 27, 2020, is used to calibrate the model initial conditions *I*(0) and infective contact rate, *β*_0_, when *β* (*t*) is taken as an explicit function of time. Other parameters are fixed at default values in Table 2. Parameter fitting was performed using nonlinear least squares algorithm im-plemented using the lsqcurvefit function in MATLAB.

#### 2.2.1 Model fitting using Mortality data only

Parameters calibrated for the model fitted for the mortality data (with *β_m_* set to be 0.1):

- The initial condition *I*(0) is fitted to be 11222.24501, that is 11223 cases should have been actually reported on March 1, 2020, but this condition seems to be high compared to the actual case data on that day.
- The infective contact rate, *β*_0_ is fitted to be 1.16562.

**Figure 2:**
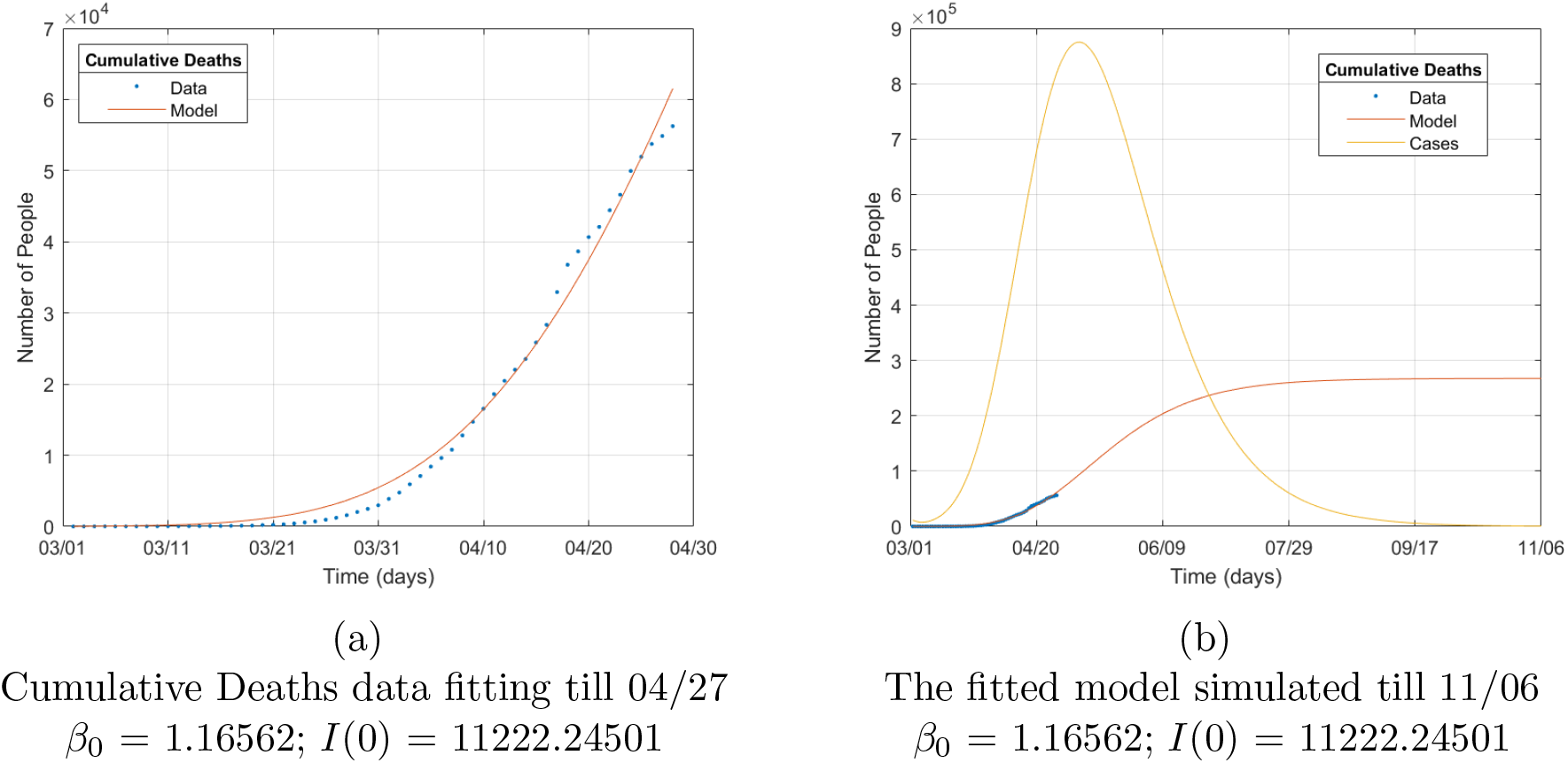
Model fitting using Mortality data only

Even though the model fits the death data nicely, the the number of cases through the time simulated is underestimated, and the initial case number *I*(0) is overestimated. Therefore, the case data should also be used while fitting the model.

#### 2.2.2 Model fitting using both Mortality and case data simultaneously

Parameters calibrated for the model fitted for mortality and case data (with *β_m_* set to be 0.1) are the initial condition and *β*_0_.

The initial condition *I*(0) is fitted to be 5848.92619, that is 5849 cases should have been actually reported on March 1, 2020, and this condition could have been the actual case data (combining the reported and unreported cases) on that day.

**Figure 3:**
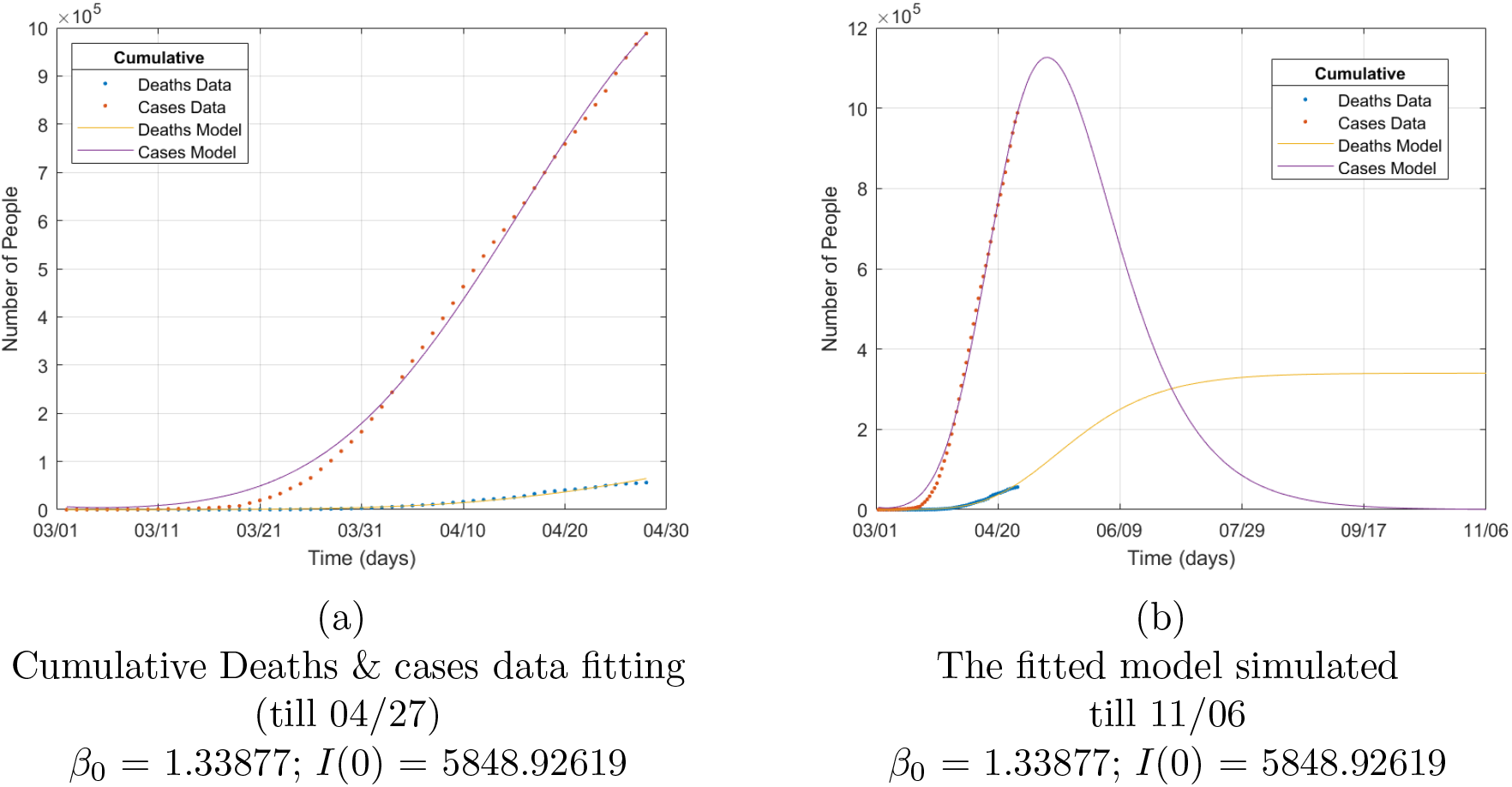
Model fitting using Mortality data only

From the fitting the data for The United States, the best fit parameters are *I*(0) = 5849; and the initial infective contact rate *β*_0_ of the time varying *β*(*t*), with fixed *r* = 0.04*day*^−1^ and *β_m_* = 0.1*day*^−1^, is found to be 1.33877*day*^−1^.

## 3 Results

### 3.1 Analytical results

#### 3.1.1 Reproduction Number (*R*_0_) & Disease Free Equilibrium (DFE)

Next generation matrices is the best way to find the basic reproduction number, beacuse it is good to follow the Infection

As seen in van den Driessche P., Watmough J. [30], *FV*^−1^ is the next generation matrix for the system at disease free equilibrium.
where,

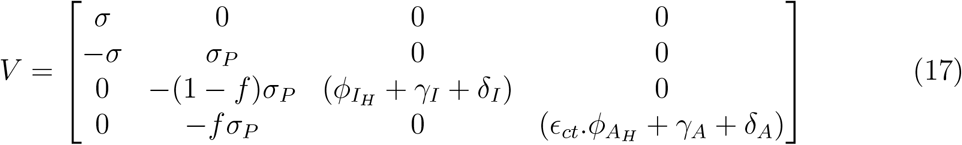

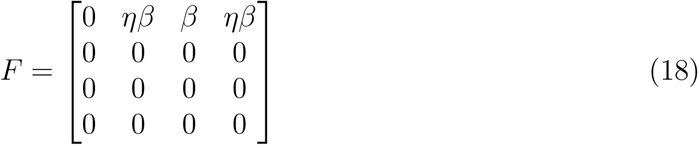

The system is global asymptotic stable (GAS) iff all eigenvalues of the next generation matrix have norm less than one, that is the spectral radius of the next generation matrix (*ρ*(*FV*^−1^)) is less than one.

In epidemiological models this spectral Radius is called as the basic reproduction number of the model (*R*_0_).

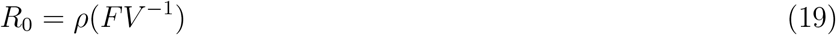

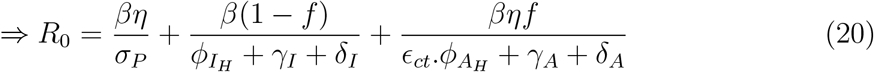

Therefore, in order to get the corona-virus to disease free equilibrium, the *R*_0_ given above should be less than 1.

Currently, the *β* value is found to be approximately 0.5. Therefore, by using all the default values given in Table 2, *R*_0_ is found to be 2.71914.

#### 3.1.2 Threshold Analysis

In the *R*_0_ equation, after taking *β* out as a common factor, it has three parts summed up together, which needs to be decreased in order to bring *R*_0_ to be less than one.

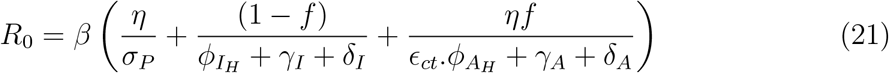

- The first part 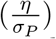 is a constant term since both *η* and *σ_P_* is fixed, and can not be controlled. using the default values this fraction is calculated to be 1.32.
- The second part 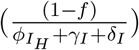, with default values for the uncontrollable variables *f*,*γ_I_*,and *δ_I_*, the only controllable variable is 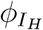 (the rate of hospitalization of the symptomatic people). To reduce the value of this whole fraction, 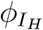 needs to be increased, by reducing the number of days it takes to admit individuals after symptom onset. This can be implemented by increasing the number of temporary hospitalization facilities in places where the cases are increasing steadily; and by increasing testing speed and availability. If the maximum possible 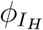 (0.033) is considered, the second part is calculated to be 2.7846.
- The third part 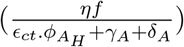, with default values for the uncontrollable variables *η*, *f*, *γ_A_*, *δ_A_*, and 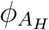, the only controllable variable is *ϵ_ct_* (the percentage of contact tracing execution). If *ϵ_ct_* is implemented at maximum capacity (0.7), the third part is calculated to be 0.5446.
- Therefore when the maximum possible rate of hospitalization is executed and contact tracing is implemented in full capacity,

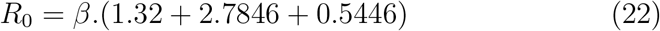

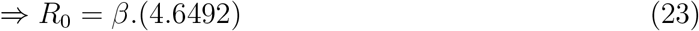

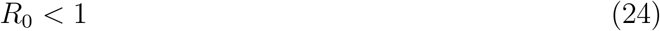

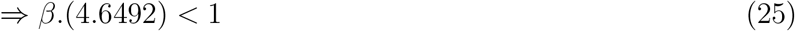

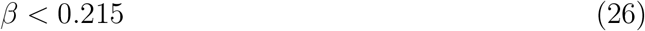 The infectious contact rate must be brought less than 0.215 to control the disease, even when the hospitalization rates are increased, and the contact tracing is implemented at full potential.

### 3.2 Uncertainity Analysis

#### 3.2.1 Uncertainity in the fraction of people remaining asymptomatic (*f*)

As presented in the report from the center of evidence based medicine [20], the percentage of people remaining asymptomatic varies drastically in many different case studies. Therefore, analyzing uncertainty in *f* is important.

As seen in the Figure 4, even 10% deviation of *f* from the default value causes a huge difference in the model simulation. It is always better to expect the worst case scenario in situations like this and make decisions so that many lives can be saved.

It can also be observed that the actual data till 4/27 lies well within the range, and the future data is expected to be within the limits till 08/28 as shown in the simulations.

**Figure 4:**
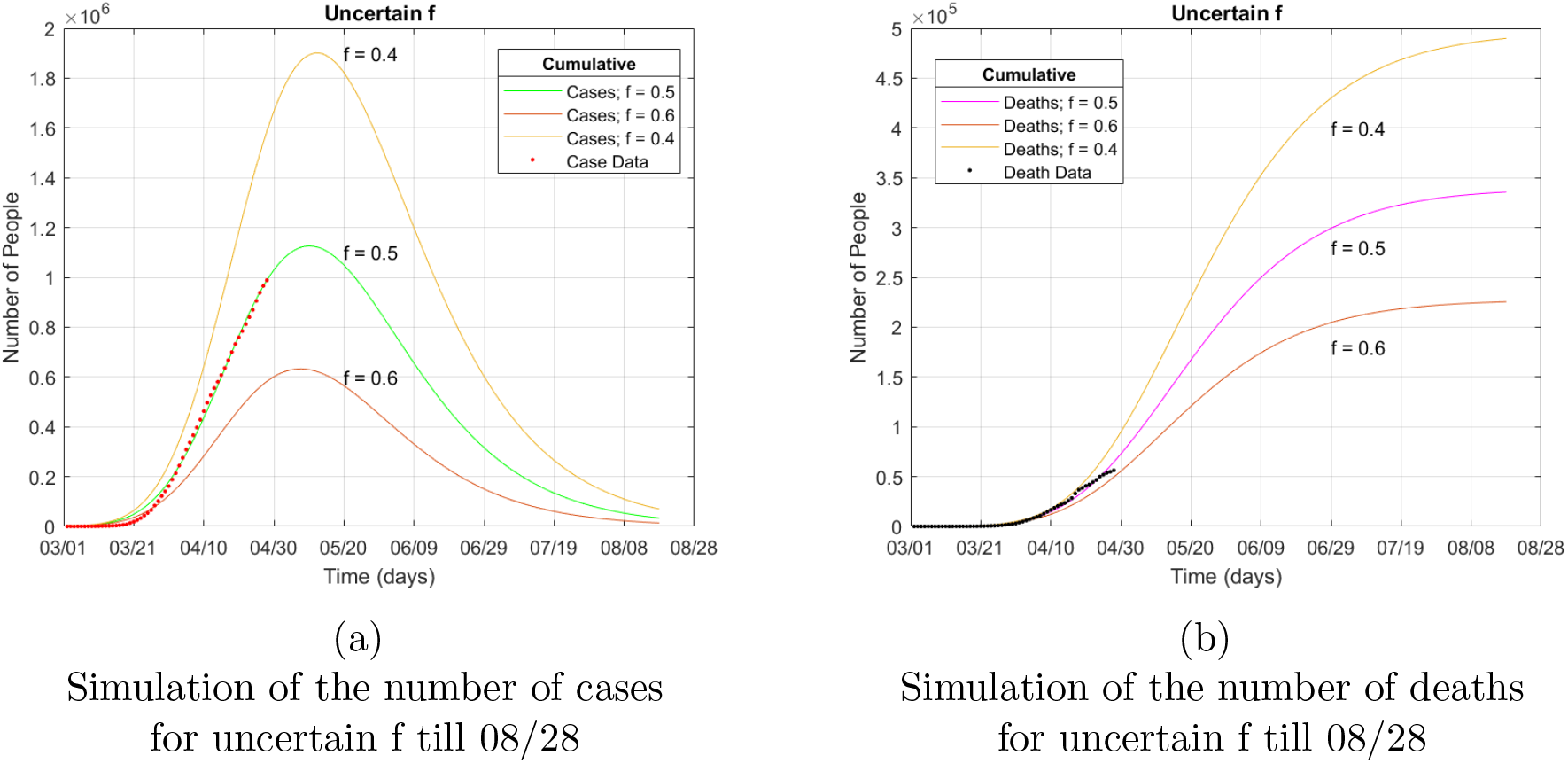
Model simulations for uncertainity in parameter *f*

#### 3.2.2 Uncertainity in infectiousness of people with no symptoms (*η*)

The infectiousness vary from person to person, this factor is very sensitive and affects the predictions hugely. Therefore, analyzing uncertainty in *η* is also important.

As seen in the Figure 5, even 4% to 6% deviation of *η* from the default value causes a huge difference in the model simulation. It is always better to expect the worst case scenario in situations like this and make decisions so that many lives can be saved.

It can also be observed that the actual data till 4/27 lies well within the range, and the future data is expected to be within the limits till 08/28 as shown in the simulations.

**Figure 5:**
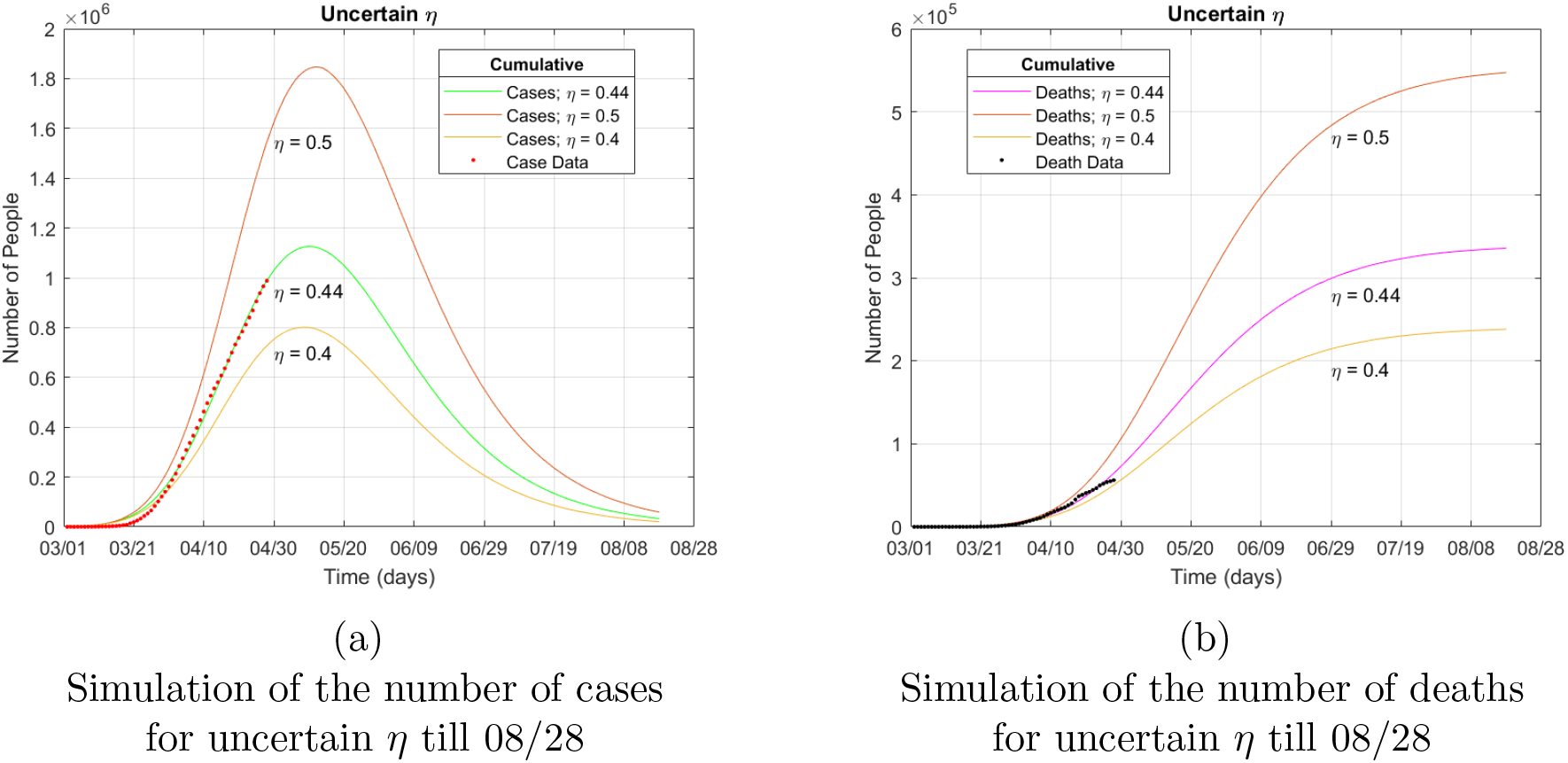
Model simulations for uncertainity in parameter *η*

### 3.3 Sensitivity Analysis

It is important to analyze how changes in control parameters of our mathematical model affects the whole model. This can be done by analyzing correlation of the parameters with Reproduction number.

- The correlation between *R*_0_ and *β* is clear from the *R*_0_ equation, they are positively correlated.

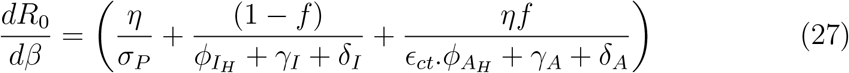 All the parameters in the equation is positive, therefore,

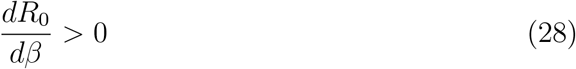 *R*_0_ and *β* has a positive correlation. When *β* is increased *R*_0_ is also increased. Therefore, to keep the reproduction number as low as possible, the infectious contact rate must be maintained as low as possible. This can be done by implementing various control strategies such as social distancing.
- The correlation between *R*_0_ and *ϵ_ct_*

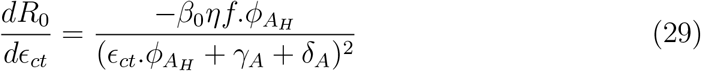 All the parameters in the equation is positive, therefore

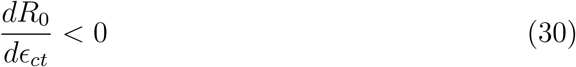 *R*_0_ and *ϵ_ct_* has a negative correlation. When *ϵ_ct_* is increased, *R*_0_ decreases. Therefore, to keep the reproduction number as low as possible, the implementation of contact tracing of Asymptomatic Individuals is crucial, and it must be maintained as high as possible.
- The correlation between *R*_0_ and 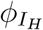

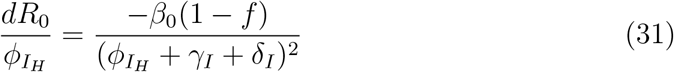 All the parameters in the equation is positive, therefore,

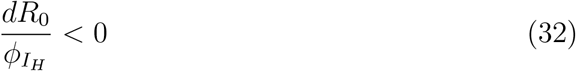 *R*_0_ and 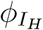 also has a negative correlation. When 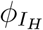 is increased, *R*_0_ decreases. Therefore, to keep the reproduction number as low as possible, the hospitalization rate should be be maintained as high as possible.
- One key difference between Isolation by contact tracing and Hospitalization, is that when people are traced, they don’t even know that they could be sick, but for hospitalization a patient need to have a severe illness.

### 3.4 Simulations of Control Strategies

Simulations of the epidemiological model is performed in MATLAB. The differential equations are solved using the function called ODE45, which solves non-stiff differential equations using medium order method. The sample code is given in the Appendix for reference

Three control strategies (Social Distancing, Contact Tracing Asymptotic population, and Increasing Hospitalization rate) are simulated, and the parameters responsible for implementing these strategies are varied and plotted together to analyze and compare. These kind of simulations helps in making decisions during a pandemic.

#### 3.4.1 Strategy 1 - Social Distancing

Strict rules on Social distancing should be implemented and followed, to prevent the spread of the disease. The simulation in Figure 6 shows that, if strategies to increase social distancing are implemented (that is, reducing *β*), the number of new cases can be reduced, and the recovery from the coronavirus pandemic is quicker. The number of estimated deaths also decreases by a significant amount in the order of 10^5^.

**Figure 6:**
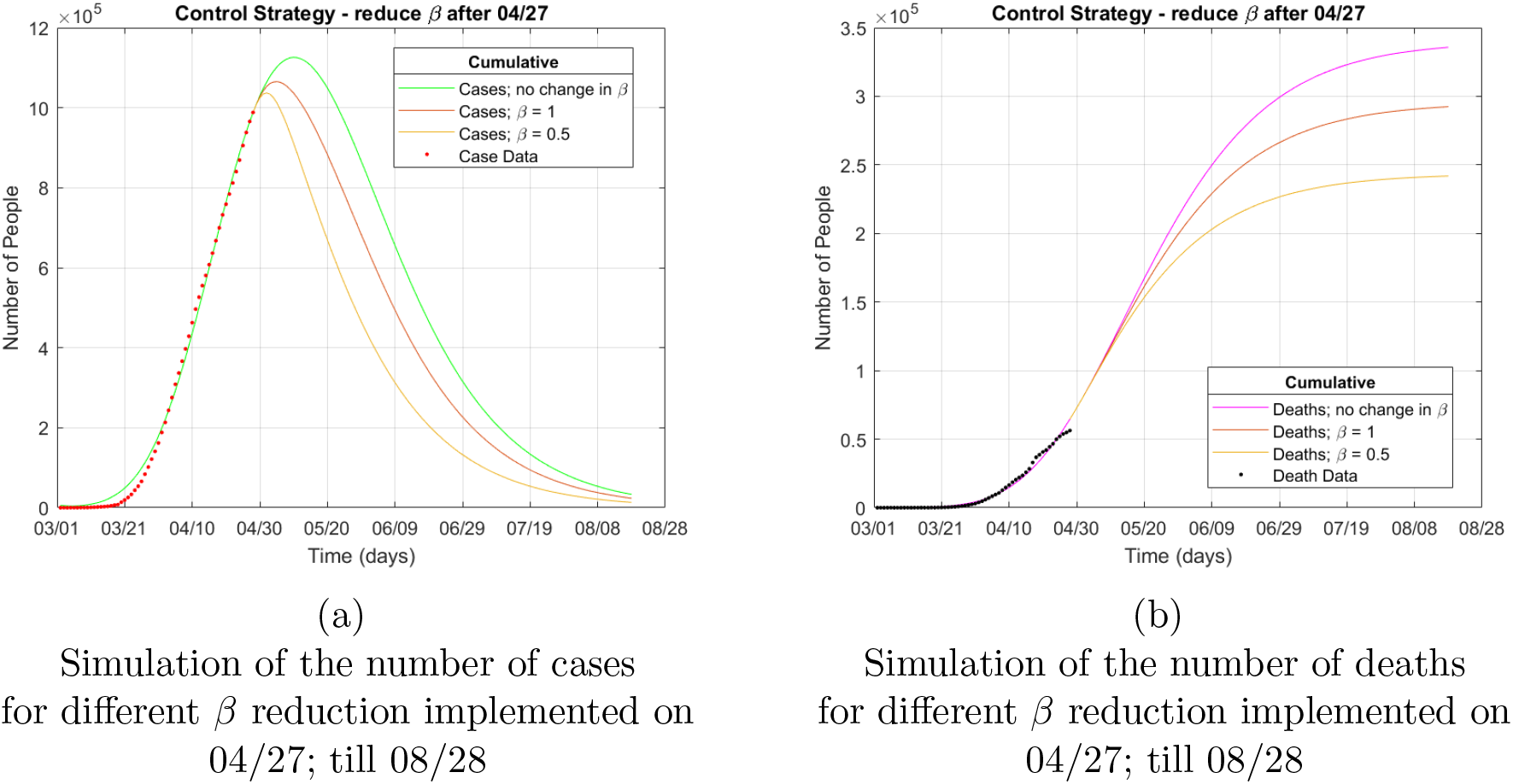
Model simulations for control strategy - Social Distancing

Two new conditions (with *β* =1 and *β* = 0.5) are simulated along with previously simulated model, and the plots are shown in Figure 6 for comparison.

The time taken to reach reasonably low number of cases is reduced by at least two weeks. Considering that the control measures are implemented continuously, the best possible time to remove social distancing without getting the disease back again, would be by early August of 2020.

With pressure from many groups of people who are affected financially, and with the government stuggling to handle the economy, the social distancing control measure is expected to be stopped in May,2020, bringing a lot of concern on how to stop the spread.

**Figure 7:**
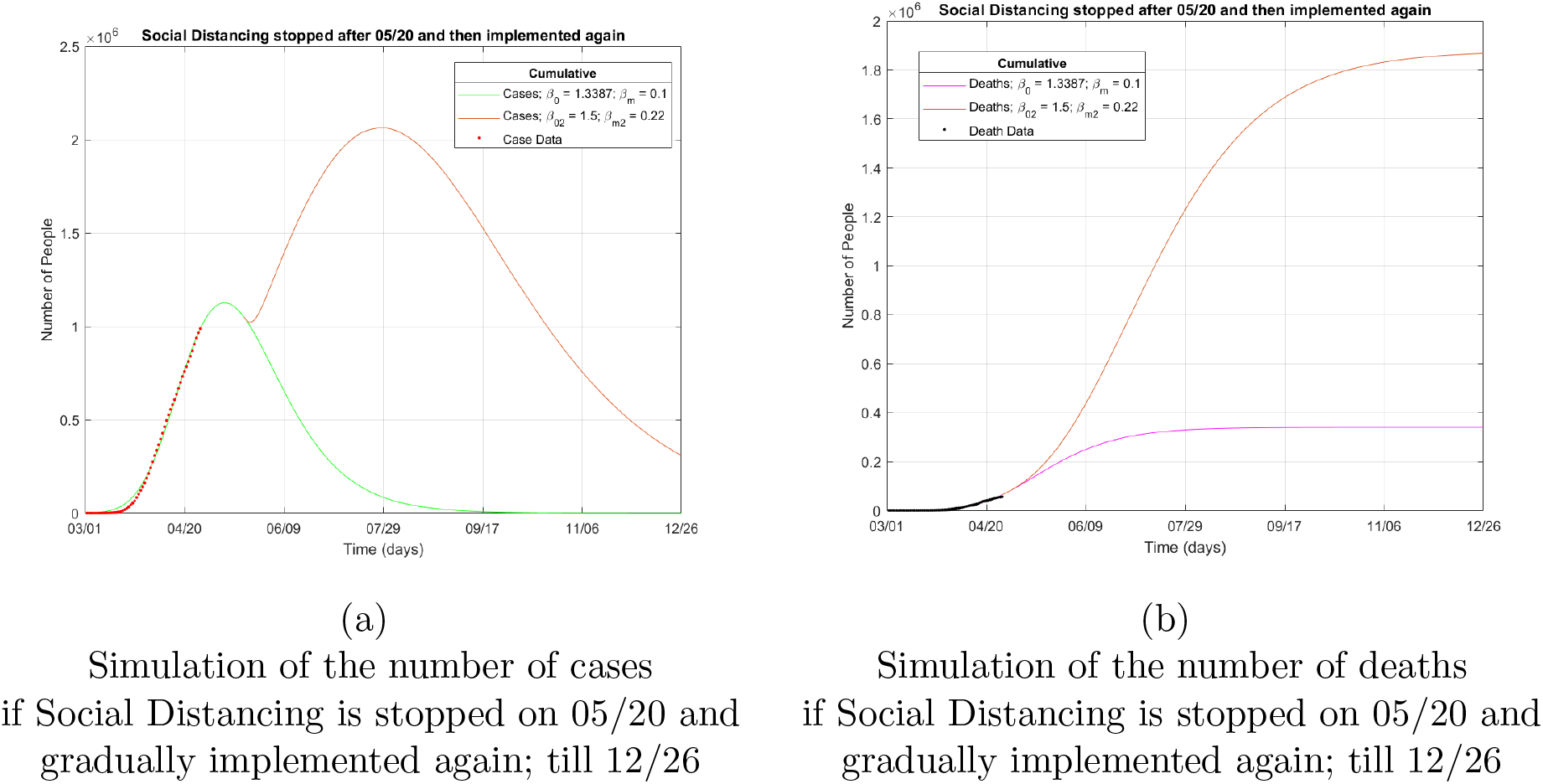
Model simulations for control strategy - Social Distancing stopped on 05/20 and followed again after noticing the increase in cases

The plot of simulations in 7 shows what will be the situation if that happens (say from 05/20). The number of cases will start to rise again, this eventually will force us to maintain social distancing again. Therefore, there will be a bigger peak in the number of reported cases, and it will take 5 more months to get out of this situation. The number of deaths will also increase as it takes longer to stop the spread of the disease.

**Figure 8:**
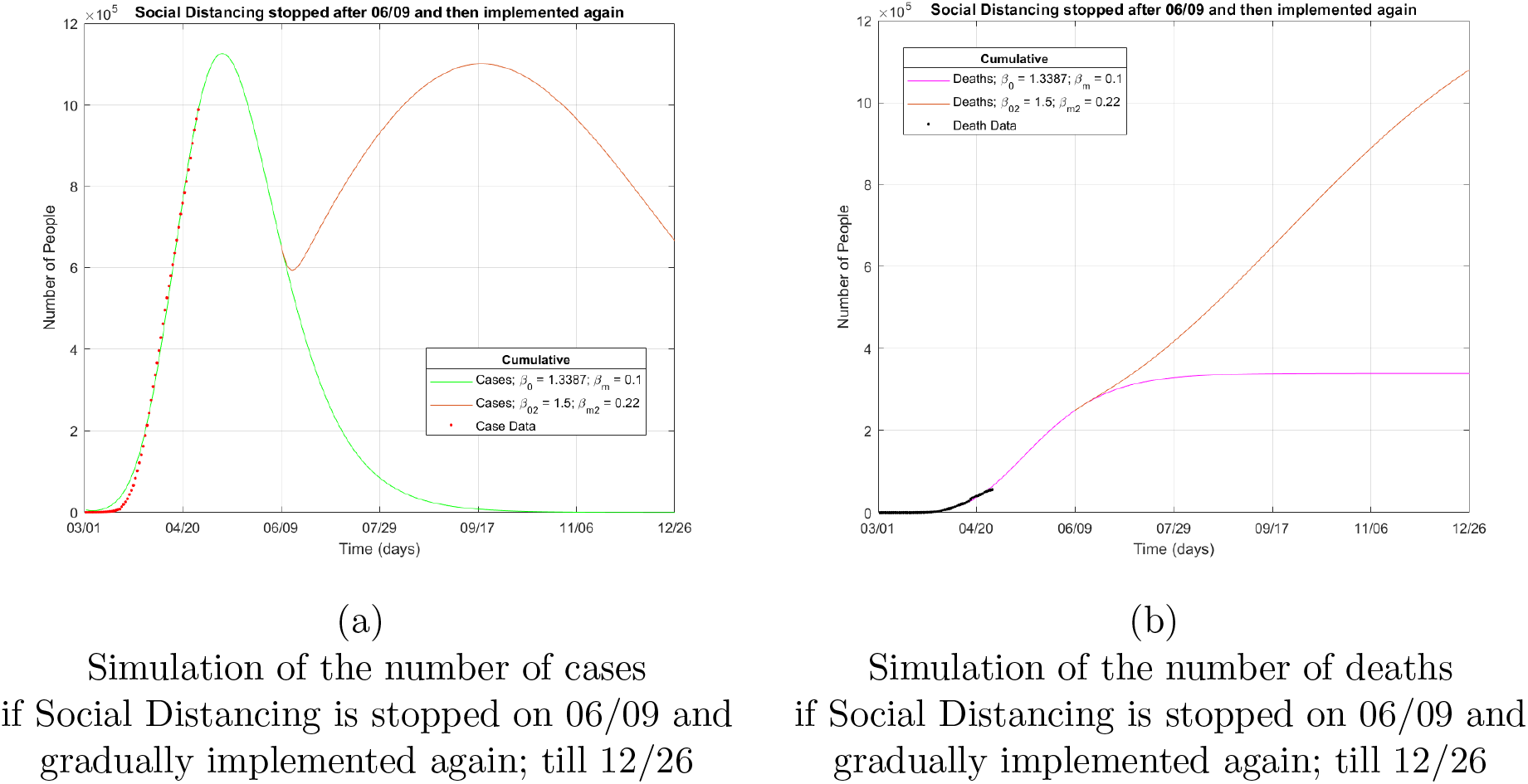
Model simulations for control strategy - Social Distancing stopped on 06/09 and followed again after noticing the increase in cases

The plot of simulations in 8 shows what will be the situation if the same thing happens from 06/09. The number of cases will start to rise again, this eventually will force us to maintain social distancing again. Even though, the peak in the number of reported cases is not increased more than what was already reached, it will take more than 8-9 months to get out of this situation. This shows us how important social distancing is.

**Figure 9:**
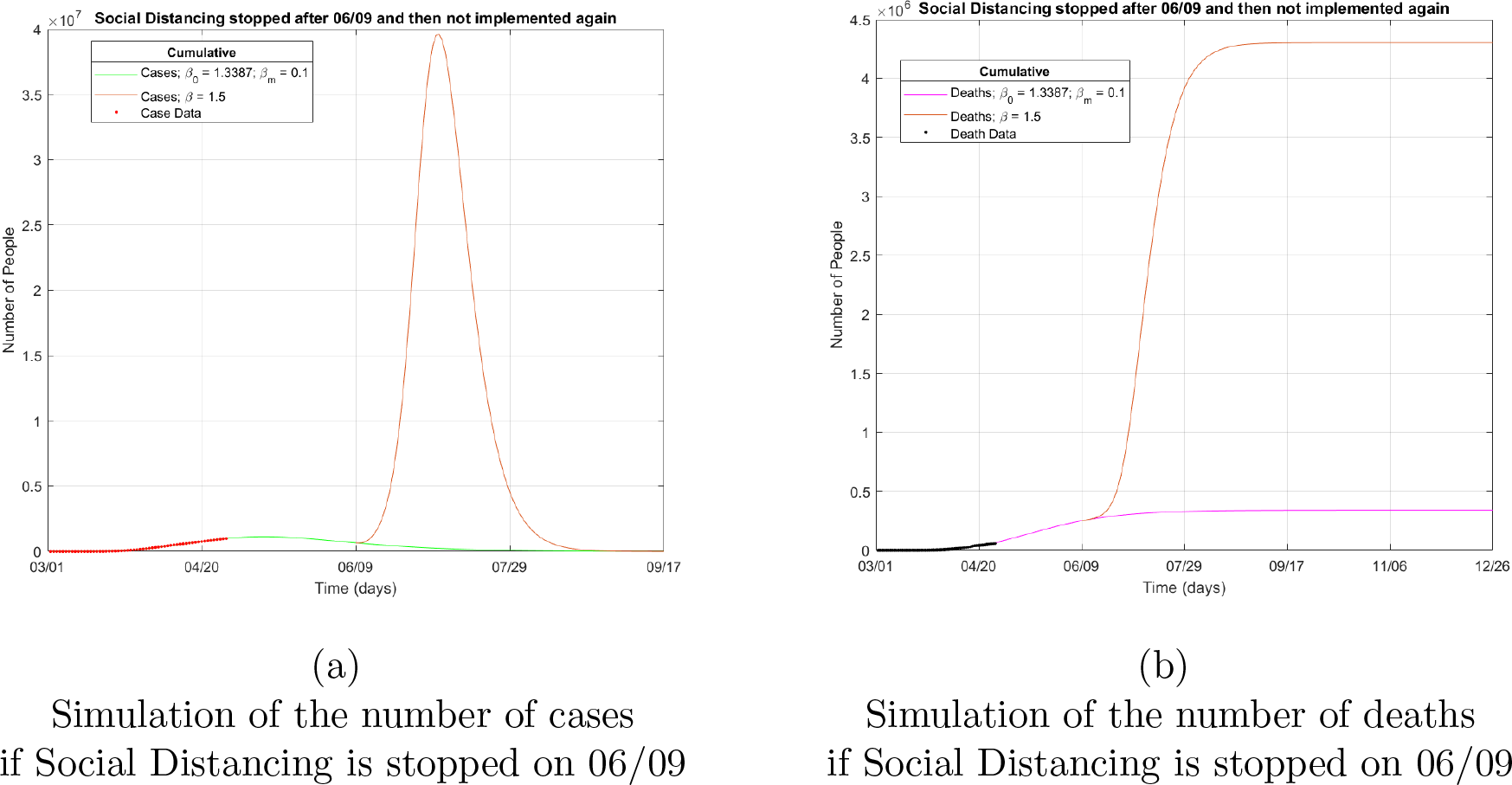
Model simulations for control strategy - Social Distancing stopped on 06/09 and not followed again

The plot of simulations in 9 shows what will be the situation if there is no social distancing at all after 06/09. The simulation is frightening as the number of cases goes upto 40 Million.

#### 3.4.2 Strategy 2 - Contact Tracing Asymptotic population

The majority of new infections are created by infected people who is showing no symptoms, they can either be totally asymptomatic or presymptomatic. To reduce the spread through them, contact tracing should be implemented. It is done by finding about all the people who were contacted by symptomatic individuals in the last two weeks. If they are tested to be positive of the disease, then they are hospitalised.

It is also possible that the person traced was the one actually who got infected, when they got in contact with this symptomatic person. Either way, contact tracing is a way to find and isolate people who could potentially carry the virus around and causing new infections. This process of isolating those who could potentially carry the disease around, is not executed at full potential, because not everyone who are exposed or asymptomatic can be found out by tracing previous contacts.

The simulations in Figure 10 shows the predictions for two new values for the percentage of contact tracing implementation (*ϵ_ct_* = 0.3 and *ϵ_ct_* = 0.7), along with the default value of 0.1. There is significant reduction in the number of cases over the month of May and after.

It can also be seen that the number of recorded death estimate increases, this is because the people who will die from this disease without knowing they had the disease, are now found and tested using contact tracing, and their deaths are recorded.

**Figure 10:**
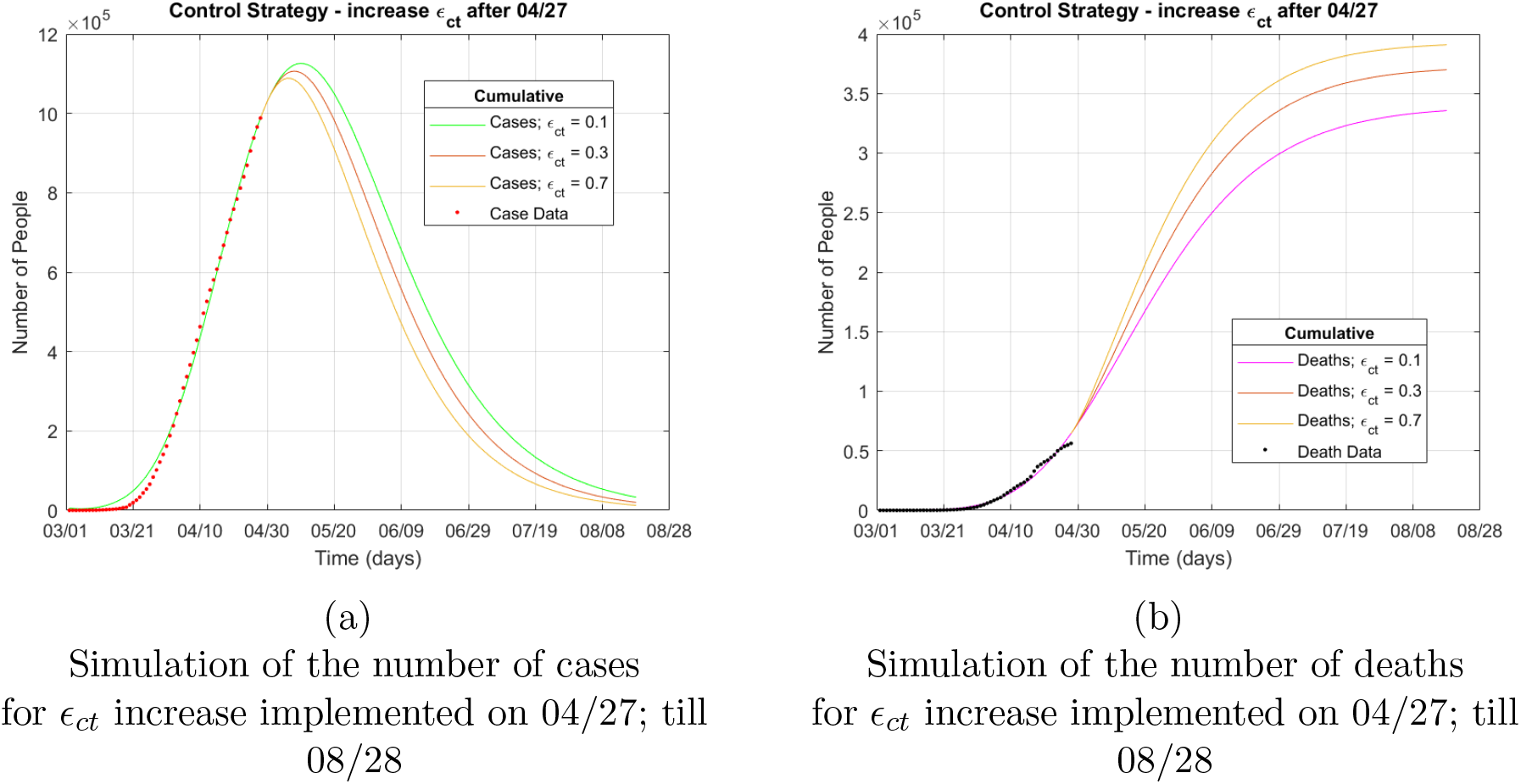
Model simulations for control strategy - Contact Tracing

#### 3.4.3 Strategy 3 - Increasing Hospitalization rate (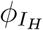)

Hospitals are struggling to handle the load of new cases. Therefore, it is reasonable to increase the number of temporary facilities and in many areas as possible. This will reduce the time taken from symptom onset to hospital admission, and thus the hospitalization increases.

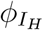 is increased in the simulations as shown in Figure 11. Two new rates - 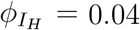 (in which takes 5 days to hospitalize someone), and 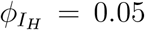 (in which takes 4 days to hospitalize someone), are simulated. It can be seen that there is a decrease in the number of reported case estimate. Increasing 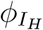 did not impact the number of cumulative deaths very much, as seen in the simulations below.

**Figure 11:**
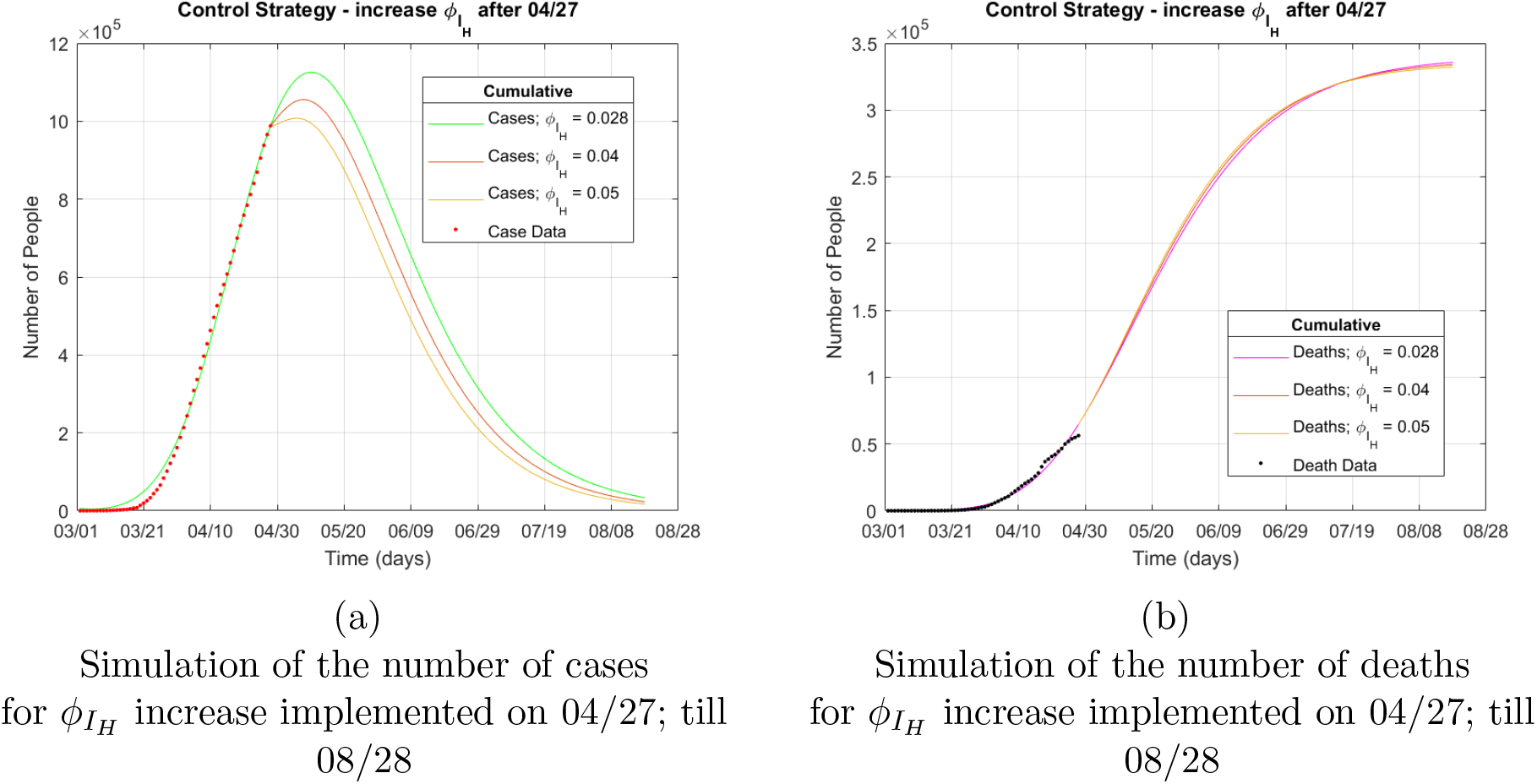
Model simulations for control strategy - Hospitalization

From the simulations of the three control strategies, Social distancing seems to have a bigger reduction in the number of diagnosed cases. No single control meothod will give the best result. It is only when all the control strategies are implemented, the considerable change is seen.

## 4 Discussion & Conclusion

A good compartmental framework of the model is designed by including all major classes of people. The most unnoticed classes during the modeling process of the coronavirus are the Pre-Symptomatic people and those that are in need of critical care. They are included in the model, and the model parameters are defined using the data from various research papers, official reports, and interviews. Once the parameters are fixed, model is fitted to the cumulative case and mortality data given by John Hopkins University. The fit gave the values for *β*_0_ to be 1.33877 and *I*(0) to be 5849, which actual infected cases in the US (on March 1, 2020).

Different aspects of the model have been analyzed and the effect of various control strategies have been simulated.

The disease free equilibrium is discussed and using the threshold analysis, basic contact rate to achieve DFE is calculated to be 0.215. The basic Reproduction Number is also calculated using the default values, and is found to be 2.71914, which is in the range similar to what most modeling papers produce.

Uncertainty in the fraction of the infectious people being asymptomatic and in the infectiousness of people with no symptoms are simulated, and the worst case scenarios are found. In the worst case, the number of cases can go up to 1.9 Million by second week of March, 2020. The death toll can go up to 0.5 Million by the end of August, 2020.

Basic sensitivity analysis is performed, and the correlation between different control parameters and the basic reproduction number are found. *β*_0_ is the positively correlated, and therefore should be reduced. The contact tracing implementation and hospitalization rate of symptomatic people are inversely correlated, and therefore they must be increased as much as possible.

The Control strategies are simulated and the results are analyzed. The Social distancing strategy seem to be best possible one right now. With the absence of vaccines, it is better to prevent the disease. If the social distancing strategy was highly implemented as shown in the simulations the number of cases during the peak can be reduced up to 1.1 Million. The duration of the implementation is also reduced by at least two weeks.

Other simulations have also been added to discuss what will happen if the social distancing is stopped in mid of May, 2020, or June, 2020 and then the control measure is followed again. If the distancing is stopped in May and implemented again, it will take till the end of the year to recover from the disease. If the distancing is stopped in June and implemented again, it will take us to first few months of next year, which is not a welcomed situation.

Another simulation in which the social distancing is stopped totally on 06/09 is plotted and the results are alarming. The peak will drastically rise up to 40 Million, and the death toll will go upto 4.5 Million. Fortunately, this will not happen, because the control measure can not be stopped abruptly as done in the simulations, and the government will take actions before something like that happens.

Another Control Strategy is contact tracing of asymptomatic individuals. The simulations shows that the number of cases can be decreases, though the change is not as predominant as it was in the case of social distancing. Anyhow, implementing contact tracing along with social distancing will produce a good improvement of the control of the disease.

The last control measure simulated was the hospitalization rate. Increasing this rate is difficult, but even minor changes can bring a huge change. The current time it takes to hospitalize someone showing symptoms is 7 days. The simulation shows the effect of reducing this time to 5 and 4 days. As the rate increases to 0.05 (that is 4 days for admission), the peak number of cases could have been reduced to 1 Million.

In summary, this paper shows the advantage of mathematical modeling and their simulations during a pandemic like COVID-19. The process helps in making good well informed decisions. In our case the United States of America should stay in lock down at least until July, 2020, to prevent major catastrophe. Despite the protest against the lock down and people’s social needs, the control strategies should be implemented for the greater good. It is only when humanity works together, this pandemic can be eradicated.

## Data Availability

The state-level time series for both cumulative mortality data and cumulative case data, compiled by Center for Systems Science and Engineering at Johns Hopkins University, from March 1, 2020, through April 27, 2020, is used to calibrate the model parameters and initial condition.

https://github.com/CSSEGISandData/COVID-19/tree/master/csse_covid_19_data/csse_covid_19_time_series

## Appendix: MATLAB Implementation of the model simulation

% c(1) S; c(2) E; c(3) P; c(4) I;

% c(5) A; c(6) H; c(7) C; c(8) R; c(9) D

col9 = The full Cumulative death data is loaded here;

% 40th entry to 97th entry corresponds to March 1,2020 to April 27,2020 respectively

column9 = col9(40:97);

colconf = The full Cumulative case data is loaded here;

colunmconf = colconf (40:97);

tf = 170;

tspan = linspace (1, tf, tf);

td = linspace (1,58,58);

y0 = [328200000 0 0 5849 0 0 0 7 1];

[t, c] = ode45(@corona, tspan, y0);

plot(t,c(:,4),’color’,’green’);hold on;

plot(t,c(:,9),’color’,’magenta’);hold on;

plot(td,colunmconf’,’.’,’MarkerEdgeColor’, ’red’);hold on;

plot(td,column9’,’.’,’MarkerEdgeColor’, ’black’);hold off;

dateaxis(’x’, 6, ’01-Mar-2020’);

grid;

title (’\beta_0’)

xlabel (’Time (days)’);

ylabel (’Number of People’);

lgd = legend(’Cases’,’Deaths’,’Case Data’,’Death Data’,’Location’,’best’);

title(lgd,’Cumulative’);

function dcdt = corona(t,c)

eta = 0.44;

beta = 1.3387;

sigma = 1/5;

sigma_P = 1/3;

f = 0.5;

gamma_I = 1/7;

gamma_A = 1/7;

gamma_H = 1/8;

gamma_C = 1/9;

phi_I_H = 0.028;

phi_H_C = 0.0533;

epsilon = 0.1;

phi_A_H = epsilon*(1/2.7);

delta_I = 0.00154;

delta_A = delta_I/2;

delta_H = 0.00425;

delta_C = 0.00485;

r = 0.04;

b_m = 0.1;

dcdt=zeros(9,1);

N = c(1)+c(2)+c(3)+c(4)+c(5)+c(6)+c(7)+c(8);

dcdt(1)= - (1/N)*(b_m+((beta-b_m)*exp(-r*t)))*c(1)*(c(4)+eta*(c(5)+c(3)));

dcdt(2)= (1/N)*(b_m+((beta-b_m)*exp(-r*t)))*c(1)*(c(4)+eta*(c(5)+c(3))) - sigma*c(2);

dcdt(3)= sigma*c(2) - sigma_P*c(3);

dcdt(4)= (1-f)*sigma_P*c(3) - phi_I_H*c(4) - gamma_I*c(4) - delta_I*c(4);

dcdt(5)= f*sigma_P*c(3) - phi_A_H*c(5) - gamma_A*c(5) - delta_A*c(5);

dcdt(6)= phi_I_H*c(4) + phi_A_H*c(5)- phi_H_C*c(6) - gamma_H*c(6) - delta_H*c(6);

dcdt(7)= phi_H_C*c(6) - gamma_C*c(7) - delta_C*c(7);

dcdt(8)= gamma_I*c(4) + gamma_A*c(5) + gamma_H*c(6) + gamma_C*c(7);

dcdt(9)= delta_H*c(6) + delta_C*c(7) + delta_I*c(4) + delta_A*c(5);

end

## Notes

### Competing Interest Statement

The authors have declared no competing interest.

### Funding Statement

No external funding was received.

